# COVID-19 Advanced Respiratory Care Educational Training Program for Healthcare Workers in Lesotho: An Observational Study

**DOI:** 10.1101/2021.10.22.21265385

**Authors:** Valerie O. Osula, Jill E. Sanders, Tafadzwa Chakare, Lucy Mapota-Masoabi, Makhoase Ranyali-Otubanjo, Bhakti Hansoti, Eric D. McCollum

## Abstract

**Objectives:** To develop and implement a ‘Low Dose, High Frequency’ (LDHF) advanced respiratory care training program for COVID-19 care in Lesotho.

**Design:** Prospective pre-post training evaluation.

**Setting:** Lesotho has limited capacity in advanced respiratory care.

**Participants:** Physicians and nurses.

**Interventions:** Due to limited participation May-September 2020 the LDHF approach was modified into a traditional one-day offsite training November 2020 that reviewed respiratory anatomy and physiology, clinical principles for conventional oxygen, heated high flow nasal cannula, and non-invasive ventilation management. Basic mechanical ventilation principles were introduced.

**Outcome measures:** Participants completed a twenty-question multiple choice examination immediately before and after the one-day training. Paired t-tests were used to evaluate the difference in average participant pre- and post-training examination scores.

**Results:** Pre- and post-training examinations were completed by 46/53 (86.7%) participants, of whom 93.4% (n=43) were nurses. The overall mean pre-training score was 44.8% (standard deviation [SD], 12.4.%). Mean scores improved by an average of 23.7 percentage points (95% confidence interval [CI] 19.7, 27.6 percentage points, p<0.001) on the post-training examination to a mean score of 68.5% (SD, 13.6%). Performance on basic and advanced respiratory categories also improved by 17.7 (95% CI: 11.6, 23.8) and 25.6 percentage points (95% CI: 20.4, 30.8) (p<0.001). Likewise, mean examination scores increased on the post-training test, compared to pre-training, for questions related to respiratory management (29.6 percentage points (95% CI: 24.1, 35.0) and physiology (17.4 percentage points (95% CI: 12.0, 22.8).

**Conclusions:** A LDHF training approach was not feasible during this relatively early period of the COVID-19 pandemic in Lesotho. Despite clear knowledge gains the modest post-training examination scores coupled with limited physician engagement suggest healthcare workers require alternative educational strategies before higher advanced care like mechanical ventilation is implementable. Conventional and high flow oxygen are better aligned with post-training healthcare worker knowledge levels and rapid implementation.

**Strengths and limitations of this study:**

- The training aimed to use a ‘Low Dose, High Frequency’ approach to improve the competence of doctors and nurses providing advanced respiratory care to severely ill COVID-19 patients during an emergency, pandemic African context with limited advanced respiratory care services.
- To provide a foundation for future implementation of invasive mechanical ventilation and more immediate application of conventional oxygen, heated high flow nasal cannula, and non-invasive ventilation, the training coupled pragmatic respiratory anatomy and physiology concepts to key clinical principles.
- Challenges in trainee participation and respiratory equipment availability necessitated modifications to the planned ‘Low Dose, High Frequency’ training strategy that reduced both the training duration and approach.
- This evaluation provides key lessons for future COVID-19 advanced respiratory care training approaches and the respiratory modalities best aligned with current healthcare worker expertise in Lesotho and likely other similar settings.

## Introduction

The SARS-CoV-2 virus causes Coronavirus disease (COVID-19).^1^ COVID-19 severity ranges across the spectrum from asymptomatic to critically ill and includes respiratory failure requiring advanced respiratory support.^2,3^ SARS-CoV-2 has claimed over four million lives worldwide with the latest surge mainly attributable to the Delta variant.^4^ Across sub-Saharan Africa COVID-19 cases and deaths also continue to escalate, stressing already fragile healthcare systems against a backdrop of limited SARS-CoV-2 vaccine access.^4^

Lesotho is a country in southern Africa with about two million people and a 49 year life expectancy.^5^ It suffers from the second highest incidence of tuberculosis and second highest prevalence of HIV globally.^6,7^ Lesotho’s health system lacks capacity in both intensive and high care hospital services and has scarce medical oxygen resources. At the onset of the pandemic the Lesotho Ministry of Health established isolation wards for COVID-19 patients at all district-level hospitals nationally and appointed two district hospitals as dedicated COVID-19 Treatment Centers. From May 2020 USAID funded Jhpiego Lesotho, an affiliate of Johns Hopkins University, to provide COVID-19 case management support to the Lesotho Ministry of Health, with a focus on capacitating healthcare workers to provide advanced respiratory care through guideline development, training, patient care supervision, human resources support, and broader technical assistance.

The challenge of delivering quality healthcare in resource-constrained low-income and middle-income countries (LMICs) like Lesotho is well known.^8-10^ A ‘Low Dose, High Frequency’ (LDHF) training approach is an established strategy that delivers shorter trainings spaced over time and is typically supplemented with practical clinical sessions at the workplace to reinforce learning, sustain changes in provider performance, and facilitate new skill acquisition.^11^ The LDHF approach has been shown to improve provider knowledge, patient management, and outcomes in LMICs and may be more feasible to deliver in healthcare settings that cannot afford to have providers engaged in traditional offsite trainings for long periods of time at the expense of depleting patient care personnel.^12^

The COVID-19 pandemic has required a rapid pivot from longstanding priorities in southern Africa like HIV and tuberculosis care towards acute respiratory treatment and related programmatic support. Given the urgent need to capacitate medical providers to manage patients with severe and critical COVID-19 we developed and implemented a LDHF healthcare worker training course to improve knowledge and skills for advanced respiratory care. The aim of this study was to describe and evaluate the effectiveness of this training program delivered during the early phases of the COVID-19 pandemic.

## Methods

### Clinical Setting

Berea Hospital is secondary hospital located approximately 30 kilometers north of the capital city of Maseru in the town of Teyateyaneng in Berea District and served as the COVID-19 treatment center for the northern region. Mafeteng Government Hospital is a regional hospital about 75 kilometers south of Maseru in the district of Mafeteng and is designated as the COVID-19 treatment center for the southern region. From May to November 2020 Berea Hospital had three inpatient wards and 50 patient beds allocated to COVID-19 care and Mafeteng Government Hospital had one inpatient ward and 20 beds for COVID-19 patients. Clinical staffing fluctuated during this period; Berea Hospital had 6-8 doctors and 30-35 nurses while Mafeteng Government Hospital had 8-9 doctors and 25-30 nurses. Doctors and nurses were all licensed and registered to provide patient care in Lesotho.

Neither Berea Hospital or Mafeteng Government Hospital offered ‘high care’ or ‘intensive care’ services. We considered ‘high care’ an area of the hospital with higher nurse to patient ratios (i.e., 1 nurse to 5-6 patients), systems for close monitoring, and the capacity to deliver more advanced respiratory treatments like conventional oxygen and heated high flow nasal cannula to severely ill patients and selected critically ill patients breathing spontaneously and generally stable. By comparison, we considered ‘intensive care’ an area of the hospital with higher nurse to patient ratios (i.e., 1 nurse to 1-2 patients), systems for continuous, invasive monitoring and the capacity to deliver life sustaining respiratory treatments like non-invasive and invasive mechanical ventilation to critically ill patients.

### ‘Low Dose, High Frequency’ Advanced Respiratory Care Educational Training Program – Overview

The educational training program (Table 1) was designed by EDM to utilize a LDHF approach to introduce new concepts and advanced respiratory care treatments during weekly one-hour sessions spaced over several months, with supervised clinical care between sessions to cement the translation of theoretical concepts to the bedside. LDHF trainings were intended to be held onsite at the treatment centers. We developed the training content based upon review of a variety of resources as well as prior anecdotal knowledge and practical experience in providing advanced respiratory care to patients in LMICs. The trainings reviewed clinically relevant respiratory anatomy and physiology as a foundation for the principles for providing COVID-19 related advanced respiratory care. The training also included both basic and selected advanced concepts, and the fundamentals of conventional oxygen delivery (i.e., “oxygen”), heated high flow nasal cannula (i.e., “high flow”), and non-invasive ventilation (NIV). Modules were designed to be linked and to introduce concepts incrementally, so each new module built upon the previous one. Trainings were to coincide with supervised bedside experiences at the treatment centers where participants would apply their knowledge to patient care and utilize oxygen, high flow, and in selected situations NIV. In addition, modules were intended to be supplemented with case discussions of COVID patients currently or recently hospitalized to highlight key clinical teaching points. Targeted cadres were physicians and nurses that provided direct care to COVID-19 patients at the treatment centers. Hospital administrators approved the delivery of the trainings at each hospital and encouraged, but did not mandate, participation. Participants were further incentivized to participate through the receipt of continuing professional development credits and food and drinks during each session. Later in the study period we did introduce a virtual option after installing monitors and equipment at each hospital, but we did not have the capacity to provide internet data and IT support to accommodate an individualized virtual option when participants were physically present at the hospital. The broader goal was to build capacity towards ‘high care’ and set the foundation for the potential future introduction of intensive care and invasive mechanical ventilation.

**Table 1.**
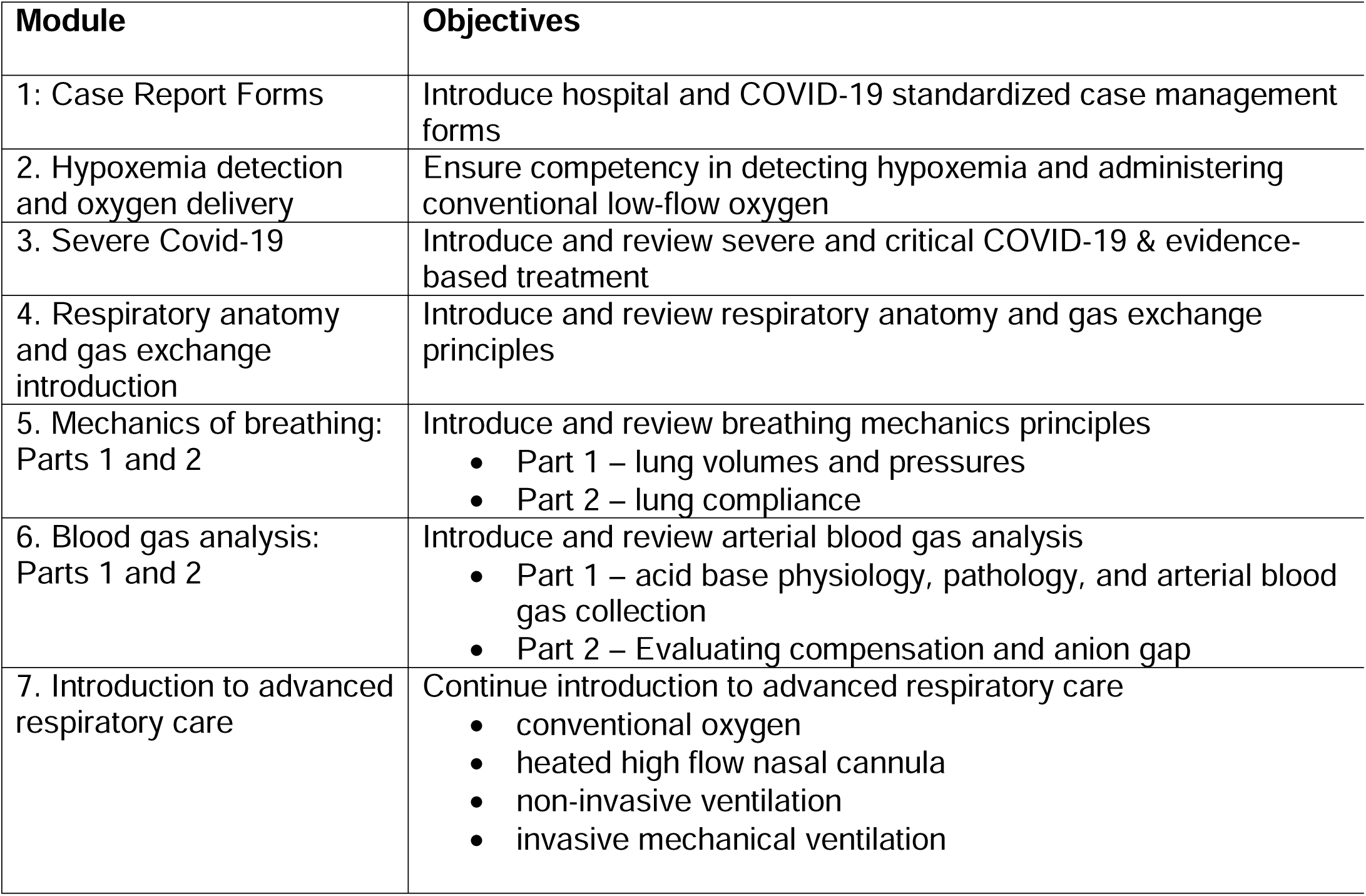
Advanced Respiratory Care Training – Modules and Primary Objectives

| Module | Objectives |
| --- | --- |
| 1: Case Report Forms | Introduce hospital and COVID-19 standardized case management forms |
| 2. Hypoxemia detection and oxygen delivery | Ensure competency in detecting hypoxemia and administering conventional low-flow oxygen |
| 3. Severe Covid-19 | Introduce and review severe and critical COVID-19 & evidence-based treatment |
| 4. Respiratory anatomy and gas exchange introduction | Introduce and review respiratory anatomy and gas exchange principles |
| 5. Mechanics of breathing: Parts 1 and 2 | Introduce and review breathing mechanics principles<br>• Part 1 – lung volumes and pressures<br>• Part 2 – lung compliance |
| 6. Blood gas analysis: Parts 1 and 2 | Introduce and review arterial blood gas analysis<br>• Part 1 – acid base physiology, pathology, and arterial blood gas collection<br>• Part 2 – Evaluating compensation and anion gap |
| 7. Introduction to advanced respiratory care | Continue introduction to advanced respiratory care<br>• conventional oxygen<br>• heated high flow nasal cannula<br>• non-invasive ventilation<br>• invasive mechanical ventilation |

### Modified Advanced Respiratory Care Educational Training Program – One-day training

After five months from May to September 2020 the LDHF approach was considered untenable due to limited and inconsistent attendance exacerbated by a lack of respiratory equipment, including high flow and NIV devices and related supplies, that had not yet been received in the country due to shipment delays. Thus, we modified the training into a one-day session to optimize participation. Content was consolidated and limited to core concepts. Due to time constraints and lack of equipment we excluded case discussions, practical hands-on sessions, and bedside patient-based teaching. All training sessions were facilitated in person by 1-2 physicians (JES, EDM). A total of 70 healthcare workers (15 physicians, 54 nurses, 1 nursing assistant) were invited to participate in any of four one-day modified trainings held between November 9 and 17, 2020 at offsite conference venues near the two treatment centers.

### Pre- and Post-training examinations of the modified one-day program

We administered a twenty question, multiple choice, paper-based examination immediately before and after the one-day training to evaluate participant baseline and acquired knowledge as an assessment of training effectiveness. All participants were requested to complete the examination individually and without training-related resources over 30 minutes. The examination included 1-3 questions per module and evaluated both basic and selected advanced topics covered in the training (see Supplemental Table 1). When appropriate to the content, questions were formulated so that learners who successfully applied their knowledge, rather than identify information by rote, would correctly answer the question.

### Ethics

The Johns Hopkins University Institutional Review Board (IRB00279223) and Lesotho National Health Research Ethics Committee (ID 12-2021) approved this research.

#### Patient and public involvement

Given the COVID-19 restrictions placed on public gatherings throughout the period of this study in Lesotho we were unable to involve and communicate to the public the development, design, recruitment, conduct, and results in this research.

### Statistical analysis

Participant performance on both the pre- and post-training examinations of the modified one-day training was assessed. The primary outcome was the average change in the overall test score between the pre- and post-test examinations. In addition, we sought to analyze if results differed by the content level (basic vs advanced) and type (respiratory physiology vs management). We used the two-proportion z-test to assess for differences in proportions. Paired *t*-tests were used to evaluate the difference in average scores of participants between the pre- and post-training examinations overall, and by content level and type. Stata (version 16.1; StataCorp, College Station, Texas) was used for all analyses.

## Results

A total of 53/70 (75.7%) invited participants attended the modified one-day training, and of the 53 attendees 46 (86.7%) completed both the pre and post-test examination and were included in this analysis. Nurses comprised 43/46 (91.3%) participants.

### Pre-training examination performance – One-day modified training

No participants achieved a score of 80% or greater on the 20 pre-training examination questions, with the lowest and highest scores of 20% (n=4) and 75% (n=15). Overall, the mean pre-training examination score was 8.9/20, or 44.8% (standard deviation (SD), 12.1%) (Figures 1 and 2, and Supplemental Table 2). The examination questions were stratified into basic (7/20, 35%) and advanced concepts (13/20, 65%). Although participants scored higher on basic topics (mean score 3.8/7 (54.0%, SD 18.8%) than advanced concepts (mean score 5.2/13 (39.8%, SD 12.6%) this difference did not reach statistical significance (p=0.542) (Figure 3 and Supplemental Table 3). Examination questions were also subdivided into respiratory physiology (10/20) and respiratory management (10/20) topics, with participants achieving average scores of 4.5/10 (45.5% (SD 18.6%)) and 4.2/10 (42.0% (SD 15.1%)) (p=0.892) (Figure 4 and Supplemental Table 4). Three doctors completed the pre-training examination and scored an average of 63.3% (SD 16.0%).

**Figure 1.**
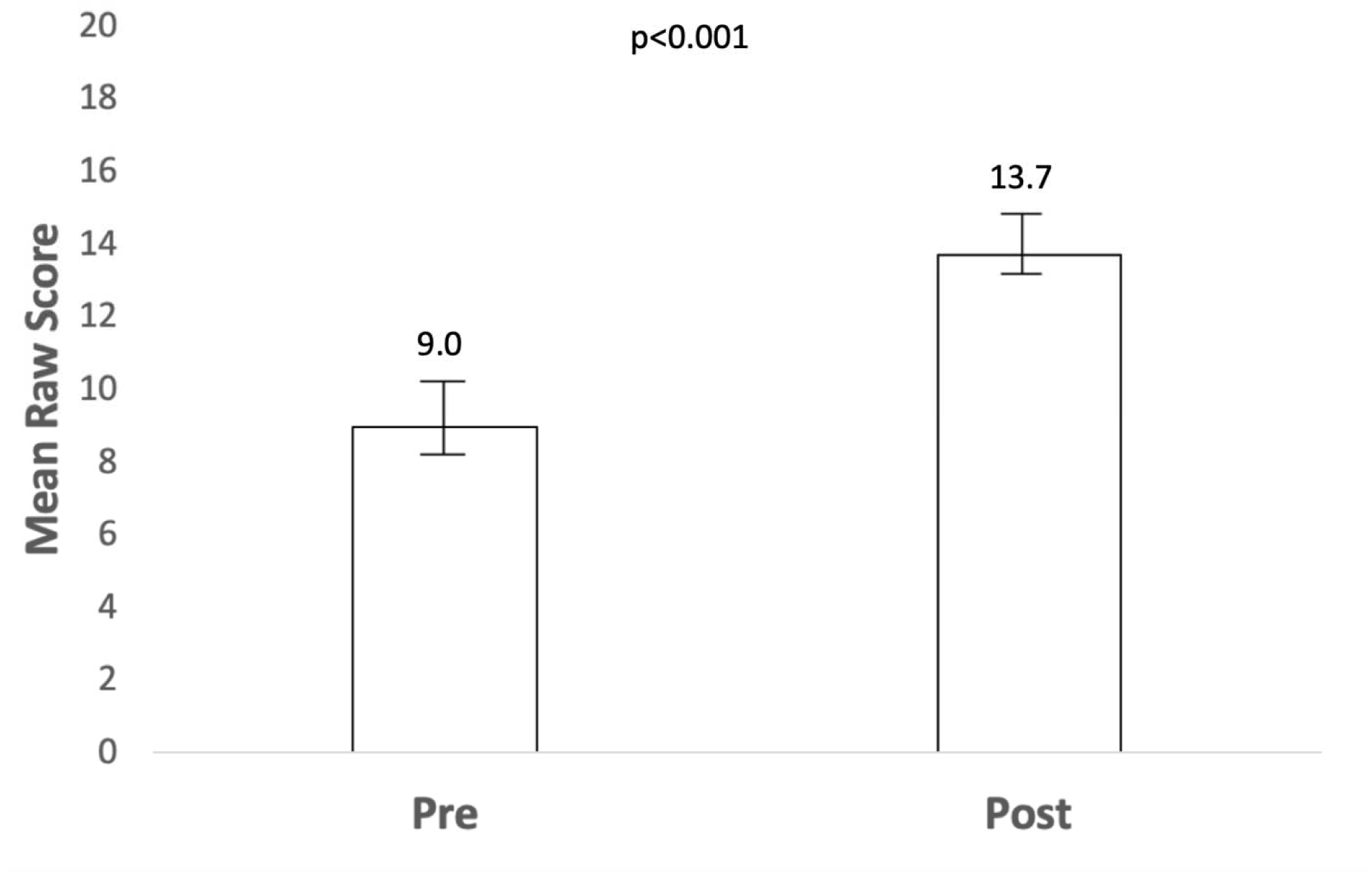
Mean pre and post-examination raw scores (out of 20). Bars represent the 95% confidence interval.

**Figure 2.**
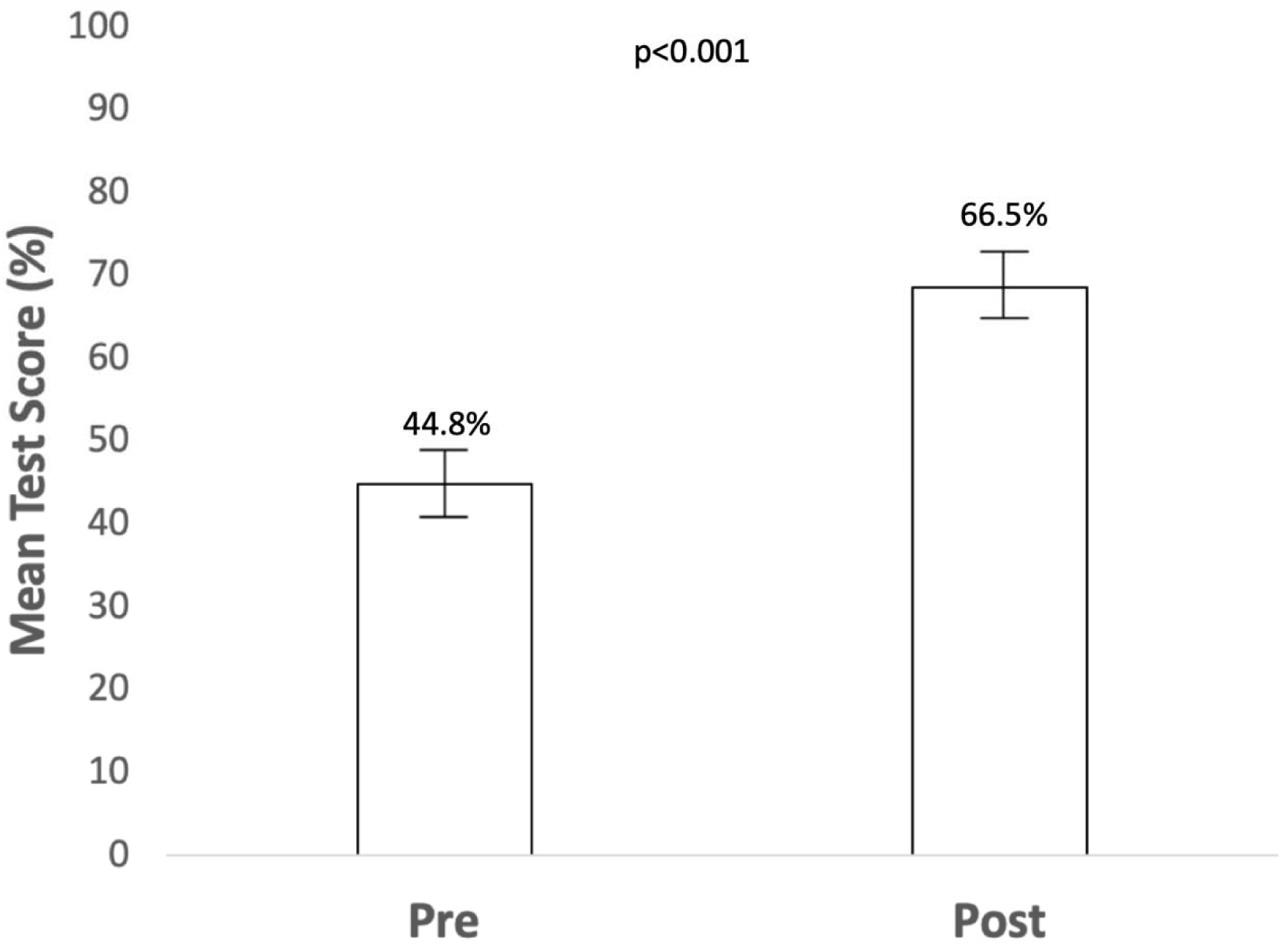
Mean pre and post-examination scores by percentage. Bars represent the 95% confidence interval.

**Figure 3.**
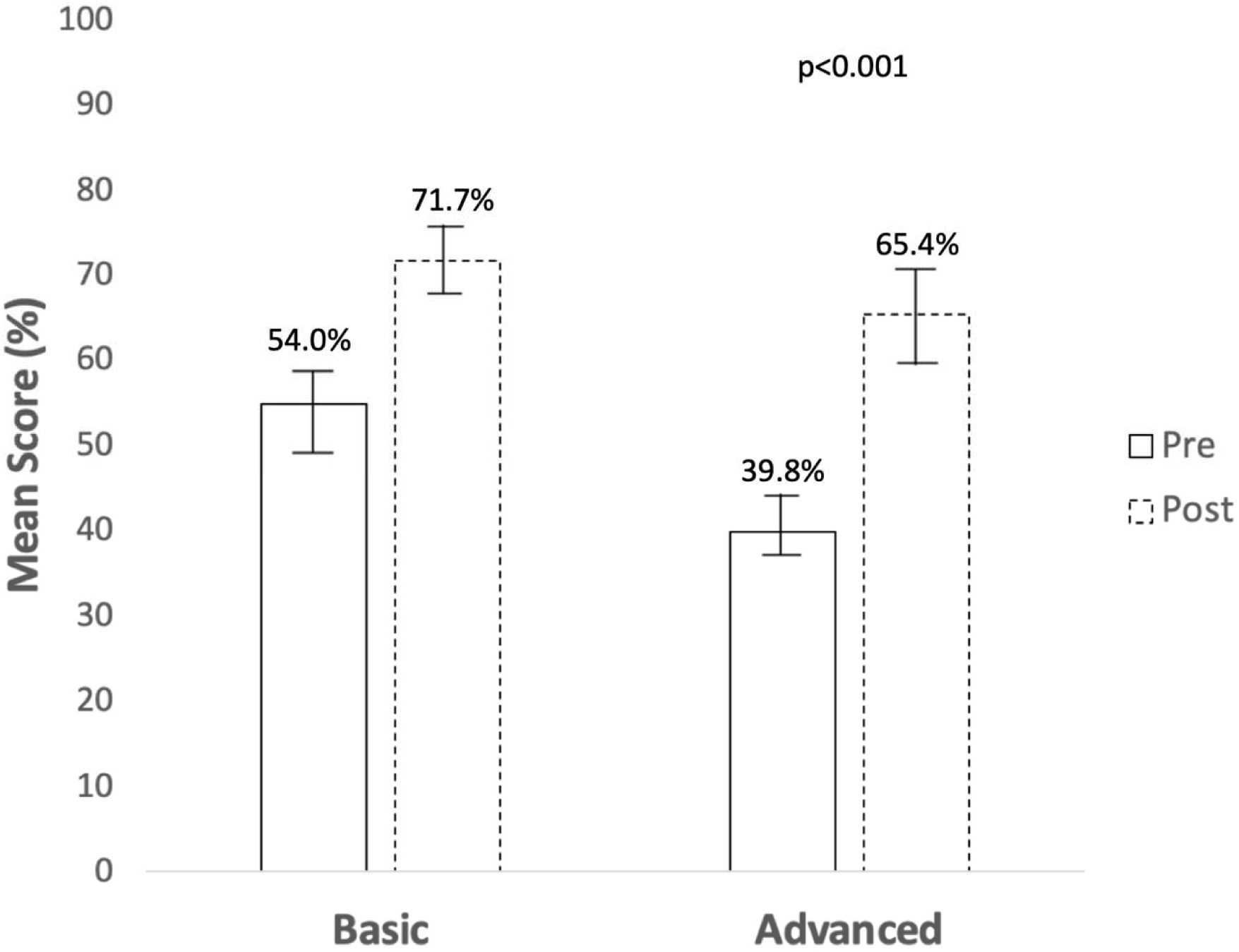
Mean pre and post-examination scores for basic and advanced concepts by percentage. Bars represent the 95% confidence interval.

**Figure 4.**
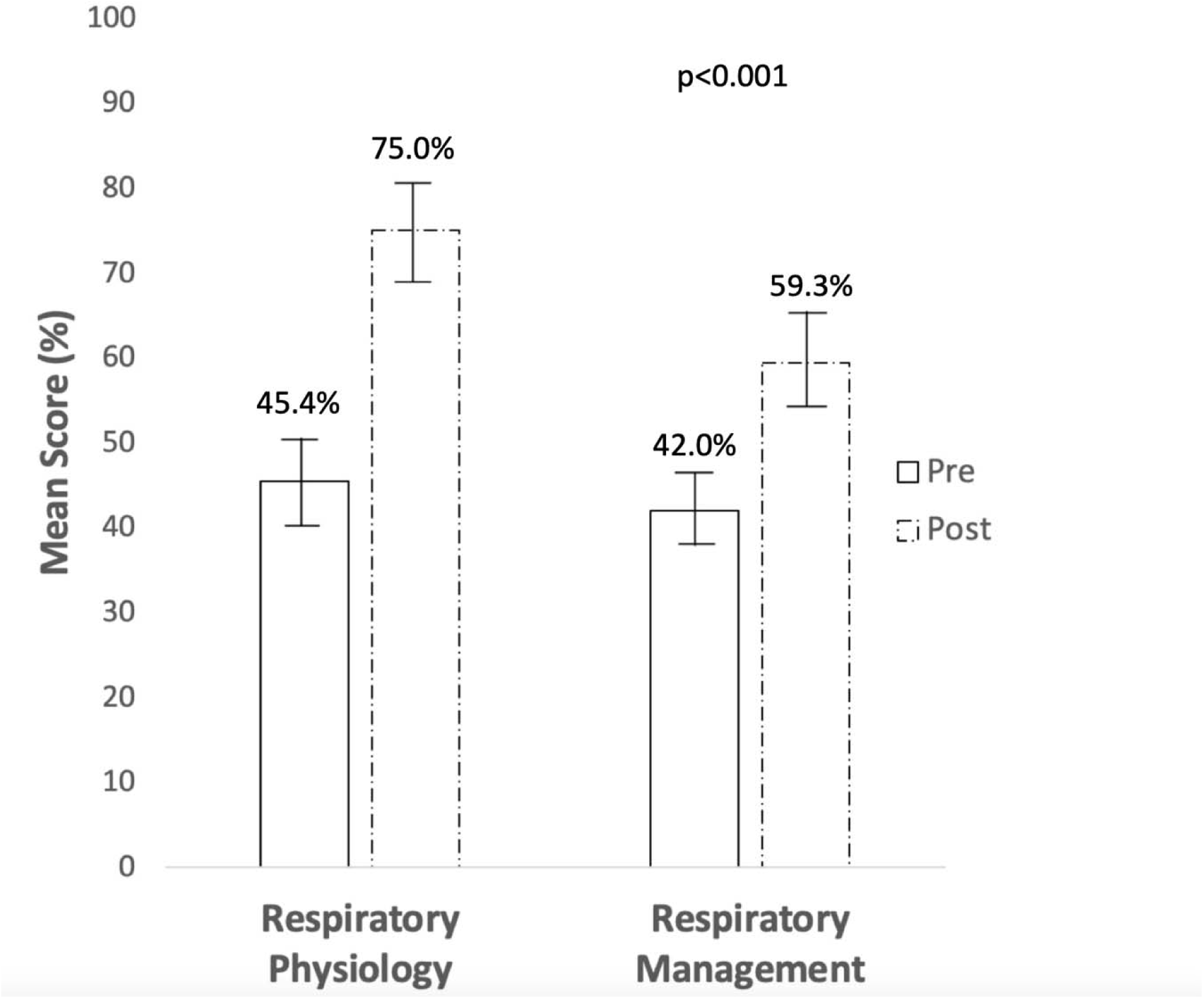
Mean pre and post-examination scores for respiratory physiology and respiratory management topic areas by percentage. Bars represent the 95% confidence interval.

### Post-training examination performance – One-day modified training

On the post training examination 9/46 (19.5%) participants scored ≥80% with an average score of 13.7/20 (68.5%, SD 13.6%) (Figures 1 and 2, and Supplemental Table 2). Two participants scored highest at 100% (n=20) and three scored lowest at 45% (n=9). The average score on basic and advanced topics was 5.0/7 (71.7%, SD 17.6%) and 8.5/13 (65.4%, SD 17.7%) (p=0.774) (Figure 3 and Supplemental Table 3) and for respiratory physiology and management was 7.5/10 (75.0%, SD 17.9%) and 5.9/10 (59.3%, SD 16.4%) (p=0.454) (Figure 4 and Supplemental Table 4). On the post-training examination doctors (n=3) scored an average of 90.0% (SD 5.0%).

### Comparing pre and post training examination performance – One-day modified training

Overall, performance improved between the pre and post-test examinations by an average of 23.7 percentage points (95% confidence interval (CI), 19.7%, 27.6%) (p<0.001) (Figure 2 and Supplemental Table 2). Similar improvements were observed for basic and advanced concepts as well as respiratory management and physiology topics (Figures 3 and 4, Supplemental Tables 3 and 4). Specifically, there was an average increase of 17.7 percentage points (95% CI, 11.6%, 23.8%, p<0.001) in basic concepts and 25.6 percentage points (95% CI, 20.4%, 30.8%, p<0.001) in advanced concepts. For respiratory management and physiology there was a mean improvement of 17.4 (95% CI, 12.0, 22.8%, p<0.001) and 29.6 percentage points (95% CI, 24.1, 35%, p<0.001) between pre and post training examinations. Amongst physicians, while there was an average increase of 26.6 percentage points (95% CI, -16.9%, 70.2%, p=0.119), this did finding did not reach statistical significance.

## Discussion

In this report we described and evaluated a new training course for advanced respiratory care administered in the sub-Saharan African country of Lesotho. The training targeted doctors and nurses responsible for treating severe to critically ill COVID-19 patients at dedicated COVID-19 treatment centers. The main goal of the training is to capacitate healthcare workers to provide advanced respiratory care for COVID-19 patients in a setting with limited acute respiratory services. We found that while average participant scores improved, both mean pre- and post-examination training scores were generally low. The two most likely reasons for these findings include overall misalignment of the training content and/or approach with participant background and experience in acute respiratory care, as well as practical training implementation challenges that may have limited its effectiveness. This training program was intended to be a LDHF educational approach delivered weekly, spaced over several months, and supported by case discussions and bedside practical training. However, we ultimately had to modify the training to a more traditional one-day offsite session to ensure full participation for all modules as healthcare providers were not consistently available week-to-week. Despite changing our approach we were still only able to engage three physicians to participate in the training. Our findings and implementation experience raise caution around how quickly and the degree to which the Lesotho healthcare system can be empowered to more immediately provide higher levels of advanced respiratory care during the COVID-19 pandemic.

Despite modifications to the LDHF training approach and modest gains in knowledge from the training, it is important to note that we did observe improvement in examination scores among nearly all participants. Overall, this shows that appropriate education and training can improve knowledge gaps, and this is consistent with prior experience.^13,14^ The three physicians that participated in the training did score well on the post-training examination, achieving an average score of 90%. While our evaluation only examined short-term knowledge retention other studies have found similar educational programs may still promote both long-term retention as well as benefit clinical outcomes.^15,16^

The generally modest participant examination scores before and after the training may reflect a mismatch between content and/or approach with trainee background and experience levels in acute respiratory care. Given the participants were all registered medical professionals they were expected to have a pre-existing working knowledge of the fundamentals of patient respiratory care. The training was additionally premised upon the notion that linking clinically relevant respiratory anatomy and physiology to key clinical principles of advanced respiratory care would build an appropriate foundation for healthcare workers to both understand and apply concepts at the bedside. Hence, the content of several modules reviewed key areas of respiratory physiology like lung pressures, volumes, compliance, and acid-base concepts that underly high flow, NIV, and invasive mechanical ventilation delivery. An introductory understanding of these principles would enable healthcare providers to select a suitable respiratory modality for the patient, program effective and safe settings for that modality, and then appropriately monitor patient responsiveness to the treatment including adverse events and clinical deterioration. The pre-training examination scores suggest that the content at baseline may be either too new or advanced for these participants. On the other hand, the post-training examination scores also imply that a one-day approach and goal of delivering intensive care with mechanical ventilation in the near term of the pandemic needs re-evaluation.

Historically, clinical guidelines and associated trainings in LMICs tend to be more rote algorithm based rather than concept driven. The World Health Organization Integrated Management of Childhood Illness guidelines were developed in the late 1990s and are a highly successful example of this approach in LMICs.^17,18^ Implementation of these guidelines over the past two decades have contributed to substantial reductions in child mortality in resource-limited settings.^19^ While revising this advanced respiratory care training from concept building to algorithmic management is a consideration, it is ultimately our view that independent thought and problem solving by healthcare workers are a prerequisite to achieving both patient safety and positive clinical outcomes in high care and intensive care settings. While rote management algorithms seem less likely to achieve both safe and successful NIV and invasive mechanical ventilation care due to the inherent clinical complexities of these modalities, conventional oxygen delivery and high flow are relatively simpler and may be better aligned with algorithmic management. Thus, from the perspective of healthcare workers, oxygen and high flow may be more feasible for LMICs like Lesotho to rapidly upscale, while NIV and invasive mechanical ventilation are likely to require longer term, more intensive, alternative educational strategies.

We also faced multiple practical challenges implementing this training and this may also have contributed to its modest impact. Initially, we planned to disseminate the training using a LDHF approach on a weekly or bi-weekly basis spaced over several months. This would allow the content to be spread out and better digested by learners and allow for an opportunity to interweave COVID-19 patient-based discussions to enrich the module content and solidify learning. In addition, we intended to have hands on practical sessions with new respiratory equipment like high flow and NIV. We attempted this approach between May and September 2020, but we were unable to consistently engage healthcare workers on or off duty and attendance was inconsistent despite participation incentives (e.g., food and refreshments and continuing professional development credits). Provider availability was also further constrained due to requirements for a two-week quarantine after clinical shifts. Although equipment was installed at each treatment center to facilitate virtual trainings and meetings, we did not have the capacity to conduct virtual trainings to individuals unable to be at the hospital as many participants lacked laptops, Wi-Fi, and/or funds for data. We also experienced lengthy delays in the arrival of respiratory equipment into the country and high flow and NIV equipment was not available in Lesotho at this time. Collectively these challenges made it difficult to build on key concepts, hold active dialogue on cases, and utilize hands on sessions to facilitate translating concepts from the theoretical to the tangible.

As such we transitioned trainings into a traditional offsite one-day session, which required a modified approach that compressed the training content, limited case-based discussions, and reduced practical hands-on experience. Given simulation training along with group problem solving can be more effective at improving performance and knowledge, future traditional trainings will be better served if done over multiple days – or as initially planned over several months – to allow more time for these key complementary approaches.^20,21^ Based on our challenges facilitating a longer more varied LDHF training approach it will be important to monitor how a longer traditional training approach impacts provider participation and costs. Further evaluation of the degree to which a traditional training approach impacts clinical outcomes of COVID-19 patients as well as longer term knowledge retention are needed. Given conditions around the pandemic have matured over the past year a LDHF approach could also be reattempted.

There are two additional limitations worth noting. First, these results primarily reflect nurses as only three doctors participated. While the backbone of clinical care is nurses and nurses are key to patient monitoring during advanced respiratory care, this training may be more suitable for doctors than nurses. Given the very limited doctor participation we were unable to stratify our analysis by cadre as initially planned. For advanced respiratory care to be successful it will be important for doctors to participate in future trainings and reasons for their lack of attendance need clarification. In addition, given the severe human resource constraints in the health sector of Lesotho, nurses need to function independently when providing advanced respiratory care since doctors are few and unable to be continuously available for all patients. Second, to deploy the training quickly we made assumptions about the baseline educational background and working medical knowledge of providers. Before revising and redeploying this training a deeper understanding of healthcare worker educational backgrounds is needed.

In sum, this study illustrates the challenges and lessons learned in designing and administering an advanced respiratory care educational training program in Lesotho during the COVID-19 pandemic. If a LDHF approach is not feasible then future renditions of this training will need to be lengthened to at least two days and better incorporate case based and simulation training with respiratory equipment. Longer term educational and training strategies for NIV and invasive mechanical ventilation that are feasible during COVID-19 in Lesotho also need development and a LDHF approach could be revisited now that the pandemic has matured, but our findings suggest these interventions are unlikely to meaningfully impact COVID-19 care in the immediate term. Conventional and high flow oxygen approaches – as well as a stronger emphasis on rote management algorithms – are likely to be a more successful short-term strategy for rapidly strengthening capacity in advance respiratory care for severe and critically ill COVID-19 patients in Lesotho.

## Data Availability

All data produced in the present study are available upon reasonable request to the authors

## Acknowledgements

We thank the healthcare workers who participated in this training and the Lesotho Ministry of Health for their support of this work.

## Author contributions

EDM was responsible for conceptualization and design. EDM curated the data. EDM and JES collected the data. EDM and VO were responsible for data analysis. EDM and VO were responsible for data interpretation. EDM and VO wrote the original draft. EDM, VO, JES, TC, LMM, MRO, BH, and AR were responsible for writing, review, and editing.

## Competing Interests Statement

EDM declares other grants from The Bill & Melinda Gates Foundation, National Institutes of Health, Pfizer, The Save the Children Fund (UK), and United States Centers for Disease Control; EDM declares consulting fees from Aurum Institute and unpaid committee participation with the Lifebox Foundation (UK) and World Health Organization. The other authors declare no competing interests.

## Funding Statement

This report was made possible with support from the United States Agency for International Development funded RISE program, under the terms of the cooperative agreement 7200AA19CA00003. The contents are the responsibility of the authors and do not necessarily reflect the views of USAID or the United States Government.

## Data availability statement

Data are available upon reasonable request. A de-identified dataset and data dictionary will be provided after a signed data access agreement with the authors.

---

**Supplemental Table 1:** Pre/Post Test Assessment Questions by Difficulty Level and Topic Area

|  | **Difficulty Level** |  |
| --- | --- | --- |
|  | <u>*Basic*</u> | <u>*Advanced*</u> |
| Questions | 1,3,5,10,17,19,20 | 2,4,6,7,8,9,11,12,13,14,15,16,18 |
|  | **Topic Area** |  |
|  | <u>*Respiratory Physiology*</u> | <u>*Respiratory Management*</u> |
| Questions | 2,6,7,8,9,10,12,13,14,18 | 1,3,4,5,11,15,16,17,19,20 |

**Supplemental Table 2.** Pre and Post-test Performance by Participant

| Participants,<br>n=46 | Cadre | Pre-test<br>Score (%)<br>N=20<sup>a</sup> | Post-test Score<br>(%)<br>N=20<sup>a</sup> | Percent<br>Difference (%) |
| --- | --- | --- | --- | --- |
| Participant 1 | Nurse | (20.0) | 75.0 | +55.0 |
| Participant 2 | Nurse | 55.0 | 75.0 | +20.0 |
| Participant 3 | Nurse | 35.0 | 50.0 | +15.0 |
| Participant 4 | Nurse | 45.0 | 75.0 | +30.0 |
| Participant 5 | Nurse | 45.0 | 55.0 | +10.0 |
| Participant 6 | Nurse | 25.0 | 60.0 | +35.0 |
| Participant 7 | Nurse<br>Assistant | 40.0 | 45.0 | +5.0 |
| Participant 8 | Nurse | 55.0 | 55.0 | 0 |
| Participant 9 | Nurse | 65.0 | 75.0 | +10.0 |
| Participant 10 | Nurse | 60.0 | 75.0 | +15.0 |
| Participant 11 | Nurse | 40.0 | 70.0 | +30.0 |
| Participant 12 | Nurse | 45.0 | 65.0 | +20.0 |
| Participant 13 | Nurse | 20.0 | 50.0 | +30.0 |
| Participant 14 | Nurse | 45.0 | 65.0 | +20.0 |
| Participant 15 | Nurse | 30.0 | 70.0 | +40.0 |
| Participant 16 | Nurse | 45.0 | 60.0 | +15.0 |
| Participant 17 | Nurse | 55.0 | 65.0 | +10.0 |
| Participant 18 | Nurse | 30.0 | 60.0 | +30.0 |
| Participant 19 | Nurse | 30.0 | 45.0 | +15.0 |
| Participant 20 | Nurse | 30.0 | 70.0 | +40.0 |
| Participant 21 | Nurse | 45.0 | 65.0 | +20.0 |
| Participant 22 | Nurse | 45.0 | 65.0 | +20.0 |
| Participant 23 | Nurse | 35.0 | 55.0 | +20.0 |
| Participant 24 | Nurse | 50.0 | 70.0 | +20.0 |
| Participant 25 | Nurse | 45.0 | 70.0 | +25.0 |
| Participant 26 | Nurse | 40.0 | 55.0 | +15.0 |
| Participant 27 | Nurse | 40.0 | 70.0 | +30.0 |
| Participant 28 | Nurse | 65.0 | 70.0 | +5.0 |
| Participant 29 | Nurse | 45.0 | 55.0 | +10.0 |
| Participant 30 | Doctor | 70.0 | 95.0 | +25.0 |
| Participant 31 | Nurse | 55.0 | 85.0 | +35.0 |
| Participant 32 | Doctor | 75.0 | 85.0 | +10.0 |
| Participant 33 | Nurse | 50.0 | 55.0 | +5.0 |
| Participant 34 | Nurse | 55.0 | 70.0 | +15.0 |
| Participant 35 | Nurse | 50.0 | 80.0 | +30.0 |
| Participant 36 | Nurse | 35.0 | 45.0 | +10.0 |
| Participant 37 | Nurse | 50.0 | 100.0 | +50.0 |
| Participant 38 | Nurse | 40.0 | 85.0 | +45.0 |
| Participant 39 | Nurse | 40.0 | 60.0 | +20.0 |
| Participant 40 | Nurse | 25.0 | 65.0 | +40.0 |
| Participant 41 | Nurse | 50.0 | 75.0 | +25.0 |
| Participant 42 | Nurse | 35.0 | 80.0 | +45.0 |
| Participant 43 | Nurse | 50.0 | 70.0 | +20.0 |
| Participant 44 | Nurse | 60.0 | 100.0 | +40.0 |
| Participant 45 | Doctor | 45.0 | 90.0 | +45.0 |
| Participant 46 | Nurse | 50.0 | 75.0 | +25.0 |
| Mean Score (SD)/mean difference (95% CI) |  | 44.8 (12.4) | 68.5(13.6) | 23.7(19.8, 27.6)* |

\*p<0.001
<sup>a</sup>Total test consisted of 20 questions

**Supplemental Table 3:** Pre and Post-test Assessment of participants by Difficulty Level

|  | **Basic** |  |  | **Advanced** |  |  |
| --- | --- | --- | --- | --- | --- | --- |
| Participants,<br>n=46 | Pre-test<br>Score<br>(%)<br>N=7<sup>a</sup> | Post-test<br>Score<br>(%)<br>N=7<sup>a</sup> | Percent<br>Difference<br>(%) | Pre-test<br>Score (%)<br>N=13<sup>b</sup> | Post-test<br>Score (%)<br>N=13<sup>b</sup> | Percent<br>Difference<br>(%) |
| Participant 1 | 14.3 | 71.4 | +57.1 | 23.1 | 76.9 | +53.8 |
| Participant 2 | 42.9 | 57.1 | +14.2 | 46.2 | 76.9 | +30.7 |
| Participant 3 | 42.9 | 57.1 | +14.2 | 30.8 | 46.2 | +15.4 |
| Participant 4 | 85.7 | 71.4 | -14.3 | 23.1 | 76.9 | +53.8 |
| Participant 5 | 57.1 | 57.1 | 0 | 38.5 | 53.9 | +15.4 |
| Participant 6 | 14.3 | 71.4 | +57.1 | 46.2 | 38.5 | -7.7 |
| Participant 7 | 57.1 | 42.9 | -14.2 | 30.8 | 46.2 | +15.4 |
| Participant 8 | 42.9 | 57.1 | +14.2 | 53.9 | 46.2 | -7.7 |
| Participant 9 | 85.7 | 85.7 | 0 | 53.9 | 69.2 | +15.3 |
| Participant 10 | 57.1 | 85.7 | +28.6 | 61.5 | 61.5 | 0 |
| Participant 11 | 42.9 | 71.4 | +28.5 | 46.2 | 61.5 | +15.3 |
| Participant 12 | 57.1 | 71.4 | +14.3 | 46.2 | 53.9 | +7.7 |
| Participant 13 | 28.6 | 71.4 | +42.8 | 23.1 | 38.5 | +15.4 |
| Participant 14 | 57.1 | 71.4 | +14.3 | 38.5 | 61.5 | +23.0 |
| Participant 15 | 57.1 | 85.7 | +28.6 | 15.4 | 53.9 | +38.5 |
| Participant 16 | 71.4 | 85.7 | +14.3 | 30.8 | 46.2 | +15.4 |
| Participant 17 | 85.7 | 71.4 | -14.3 | 38.5 | 61.5 | +23 |
| Participant 18 | 42.9 | 71.4 | +28.5 | 23.1 | 53.9 | +30.8 |
| Participant 19 | 57.1 | 57.1 | 0 | 15.4 | 38.5 | +23.1 |
| Participant 20 | 28.6 | 71.4 | +42.8 | 30.1 | 69.2 | +39.1 |
| Participant 21 | 57.1 | 42.9 | -14.2 | 46.2 | 76.9 | +30.7 |
| Participant 22 | 42.9 | 85.7 | +42.8 | 38.5 | 53.9 | +15.4 |
| Participant 23 | 42.9 | 57.1 | +14.2 | 30.1 | 53.9 | +23.8 |
| Participant 24 | 57.1 | 85.7 | +28.6 | 46.2 | 61.5 | +15.3 |
| Participant 25 | 71.4 | 71.4 | 0 | 30.8 | 69.2 | +38.4 |
| Participant 26 | 42.9 | 57.1 | +14.2 | 38.5 | 53.9 | +15.4 |
| Participant 27 | 28.6 | 71.4 | +42.8 | 46.2 | 69.2 | +23.0 |
| Participant 28 | 85.7 | 85.7 | 0 | 46.2 | 61.5 | +15.3 |
| Participant 29 | 57.1 | 57.1 | 0 | 38.5 | 53.9 | +15.4 |
| Participant 30 | 85.7 | 85.7 | 0 | 61.5 | 100.0 | +38.5 |
| Participant 31 | 42.9 | 71.4 | +28.5 | 61.5 | 92.3 | +30.8 |
| Participant 32 | 85.7 | 71.4 | -14.3 | 69.2 | 100.0 | +30.8 |
| Participant 33 | 42.9 | 85.7 | +42.8 | 53.9 | 38.5 | -15.4 |
| Participant 34 | 71.4 | 71.4 | 0 | 30.8 | 69.2 | -24.6 |
| Participant 35 | 57.1 | 57.1 | 0 | 46.2 | 84.6 | +38.4 |
| Participant 36 | 28.6 | 42.9 | +14.3 | 38.5 | 46.2 | +7.7 |
| Participant 37 | 71.4 | 100.0 | +28.6 | 46.2 | 100.0 | +53.8 |
| Participant 38 | 57.1 | 71.4 | +14.3 | 30.8 | 92.3 | +61.5 |
| Participant 39 | 42.9 | 57.1 | +14.2 | 38.5 | 61.5 | +23.0 |
| Participant 40 | 42.9 | 57.1 | +14.2 | 15.4 | 69.2 | +53.8 |
| Participant 41 | 57.1 | 85.7 | +28.6 | 46.2 | 69.2 | +23.0 |
| Participant 42 | 28.6 | 100.0 | +71.4 | 38.5 | 69.2 | +30.7 |
| Participant 43 | 57.1 | 85.7 | +28.6 | 46.2 | 61.5 | +15.3 |
| Participant 44 | 71.4 | 100.0 | +28.6 | 53.9 | 100.0 | +46.1 |
| Participant 45 | 57.1 | 85.7 | +28.6 | 38.5 | 92.3 | +53.8 |
| Participant 46 | 71.4 | 71.4 | 0 | 38.5 | 76.9 | +38.4 |
| Mean Score (SD)/mean difference (95% CI) | 54.0(18.8) | 71.7(14.6) | 17.7 (11.6, 23.8)* | 39.8(12.6) | 65.4(17.7) | 25.6(20.4, 30.8)* |

\*p<0.001
<sup>a</sup>Basic concepts were covered by 7 out of the 20 total questions
<sup>b</sup>Advanced concepts were covered by 13 out of the 20 total questions

**Supplemental Table 4:**
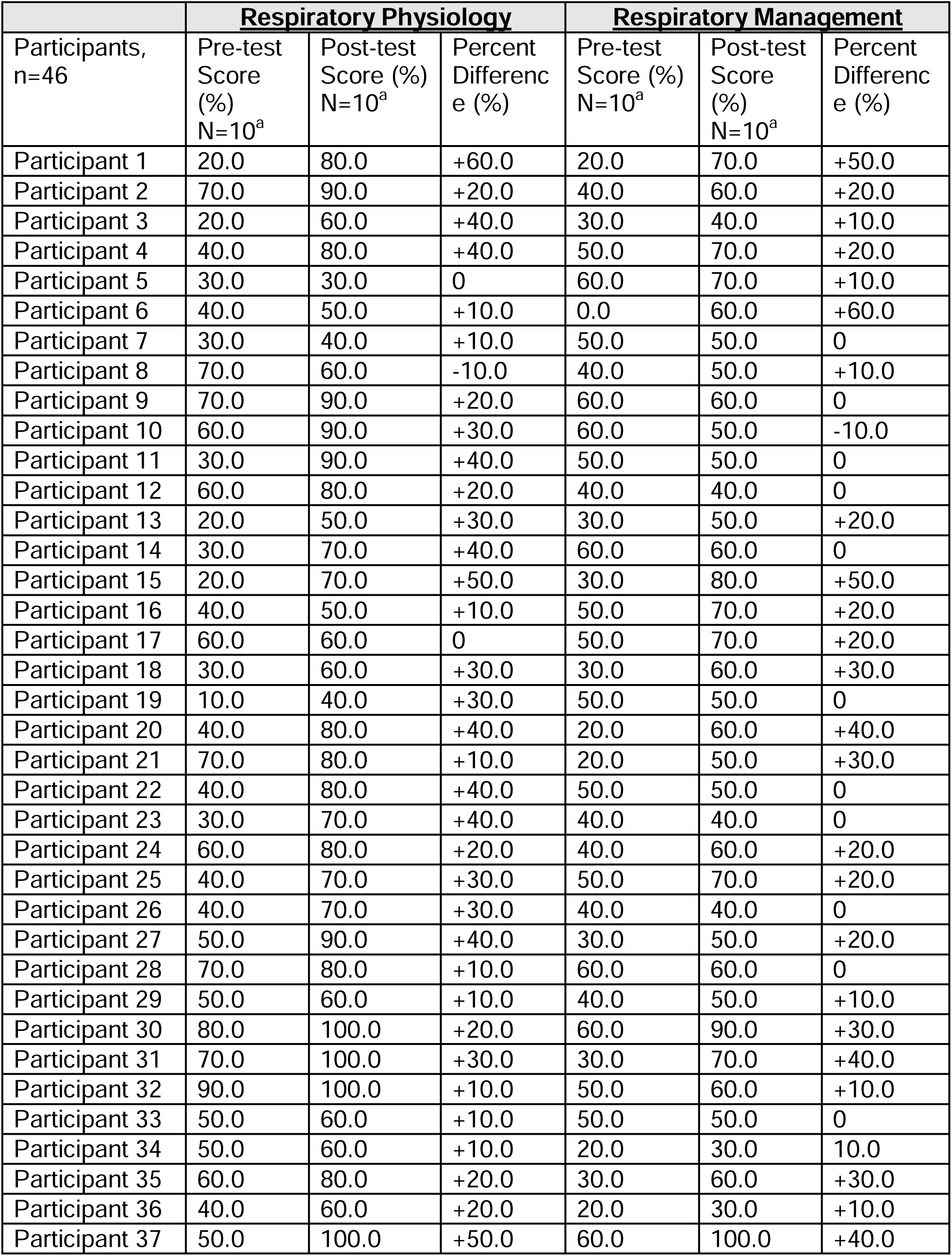
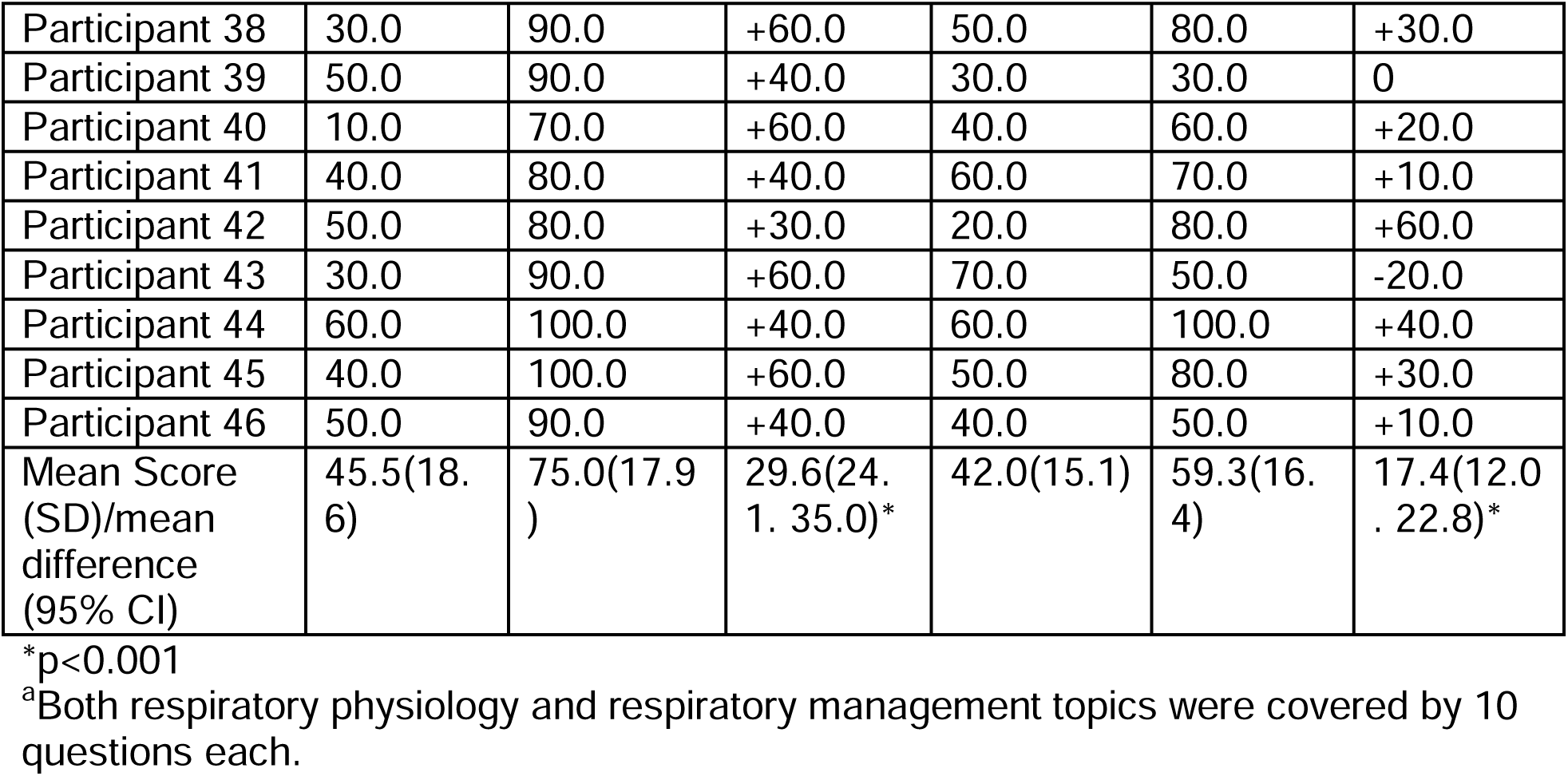
Pre and Post-test Assessment of participants by Topic Area

| Participants,<br>n=46 | **Respiratory Physiology** |  |  | **Respiratory Management** |  |  |
| --- | --- | --- | --- | --- | --- | --- |
|  | Pre-test<br>Score<br>(%)<br>N=10<sup>a</sup> | Post-test<br>Score (%)<br>N=10<sup>a</sup> | Percent<br>Differenc<br>e (%) | Pre-test<br>Score (%)<br>N=10<sup>a</sup> | Post-test<br>Score<br>(%)<br>N=10<sup>a</sup> | Percent<br>Differenc<br>e (%) |
| Participant 1 | 20.0 | 80.0 | +60.0 | 20.0 | 70.0 | +50.0 |
| Participant 2 | 70.0 | 90.0 | +20.0 | 40.0 | 60.0 | +20.0 |
| Participant 3 | 20.0 | 60.0 | +40.0 | 30.0 | 40.0 | +10.0 |
| Participant 4 | 40.0 | 80.0 | +40.0 | 50.0 | 70.0 | +20.0 |
| Participant 5 | 30.0 | 30.0 | 0 | 60.0 | 70.0 | +10.0 |
| Participant 6 | 40.0 | 50.0 | +10.0 | 0.0 | 60.0 | +60.0 |
| Participant 7 | 30.0 | 40.0 | +10.0 | 50.0 | 50.0 | 0 |
| Participant 8 | 70.0 | 60.0 | -10.0 | 40.0 | 50.0 | +10.0 |
| Participant 9 | 70.0 | 90.0 | +20.0 | 60.0 | 60.0 | 0 |
| Participant 10 | 60.0 | 90.0 | +30.0 | 60.0 | 50.0 | -10.0 |
| Participant 11 | 30.0 | 90.0 | +40.0 | 50.0 | 50.0 | 0 |
| Participant 12 | 60.0 | 80.0 | +20.0 | 40.0 | 40.0 | 0 |
| Participant 13 | 20.0 | 50.0 | +30.0 | 30.0 | 50.0 | +20.0 |
| Participant 14 | 30.0 | 70.0 | +40.0 | 60.0 | 60.0 | 0 |
| Participant 15 | 20.0 | 70.0 | +50.0 | 30.0 | 80.0 | +50.0 |
| Participant 16 | 40.0 | 50.0 | +10.0 | 50.0 | 70.0 | +20.0 |
| Participant 17 | 60.0 | 60.0 | 0 | 50.0 | 70.0 | +20.0 |
| Participant 18 | 30.0 | 60.0 | +30.0 | 30.0 | 60.0 | +30.0 |
| Participant 19 | 10.0 | 40.0 | +30.0 | 50.0 | 50.0 | 0 |
| Participant 20 | 40.0 | 80.0 | +40.0 | 20.0 | 60.0 | +40.0 |
| Participant 21 | 70.0 | 80.0 | +10.0 | 20.0 | 50.0 | +30.0 |
| Participant 22 | 40.0 | 80.0 | +40.0 | 50.0 | 50.0 | 0 |
| Participant 23 | 30.0 | 70.0 | +40.0 | 40.0 | 40.0 | 0 |
| Participant 24 | 60.0 | 80.0 | +20.0 | 40.0 | 60.0 | +20.0 |
| Participant 25 | 40.0 | 70.0 | +30.0 | 50.0 | 70.0 | +20.0 |
| Participant 26 | 40.0 | 70.0 | +30.0 | 40.0 | 40.0 | 0 |
| Participant 27 | 50.0 | 90.0 | +40.0 | 30.0 | 50.0 | +20.0 |
| Participant 28 | 70.0 | 80.0 | +10.0 | 60.0 | 60.0 | 0 |
| Participant 29 | 50.0 | 60.0 | +10.0 | 40.0 | 50.0 | +10.0 |
| Participant 30 | 80.0 | 100.0 | +20.0 | 60.0 | 90.0 | +30.0 |
| Participant 31 | 70.0 | 100.0 | +30.0 | 30.0 | 70.0 | +40.0 |
| Participant 32 | 90.0 | 100.0 | +10.0 | 50.0 | 60.0 | +10.0 |
| Participant 33 | 50.0 | 60.0 | +10.0 | 50.0 | 50.0 | 0 |
| Participant 34 | 50.0 | 60.0 | +10.0 | 20.0 | 30.0 | 10.0 |
| Participant 35 | 60.0 | 80.0 | +20.0 | 30.0 | 60.0 | +30.0 |
| Participant 36 | 40.0 | 60.0 | +20.0 | 20.0 | 30.0 | +10.0 |
| Participant 37 | 50.0 | 100.0 | +50.0 | 60.0 | 100.0 | +40.0 |
| Participant 38 | 30.0 | 90.0 | +60.0 | 50.0 | 80.0 | +30.0 |
| Participant 39 | 50.0 | 90.0 | +40.0 | 30.0 | 30.0 | 0 |
| Participant 40 | 10.0 | 70.0 | +60.0 | 40.0 | 60.0 | +20.0 |
| Participant 41 | 40.0 | 80.0 | +40.0 | 60.0 | 70.0 | +10.0 |
| Participant 42 | 50.0 | 80.0 | +30.0 | 20.0 | 80.0 | +60.0 |
| Participant 43 | 30.0 | 90.0 | +60.0 | 70.0 | 50.0 | -20.0 |
| Participant 44 | 60.0 | 100.0 | +40.0 | 60.0 | 100.0 | +40.0 |
| Participant 45 | 40.0 | 100.0 | +60.0 | 50.0 | 80.0 | +30.0 |
| Participant 46 | 50.0 | 90.0 | +40.0 | 40.0 | 50.0 | +10.0 |
| Mean Score (SD)/mean difference (95% CI) | 45.5(18.6) | 75.0(17.9) | 29.6(24.1, 35.0)* | 42.0(15.1) | 59.3(16.4) | 17.4(12.0, 22.8)* |

\*p<0.001
<sup>a</sup>Both respiratory physiology and respiratory management topics were covered by 10 questions each.

## Examination

1. **What is the oxygen saturation threshold <u>below</u> which health care providers should start low flow oxygen for COVID-19 suspect or confirmed cases? Select one answer**.
  a. <98%
  b. <94%
  c. <90%
  d. <85%
2. **Please choose the one best answer below. Patients with COVID-19 and severe ARDS have:**
  a. low lung compliance
  b. high lung compliance
  c. normal lung compliance
  d. no lung compliance
3. **Please choose the one best answer below. The treatment currently recommended for severe or critical COVID-19 patients is the following:**
  a. Hydroxychloroquine
  b. Dexamethasone
  c. Supportive care (respiratory and general care)
  d. a & b
  e. b & c
  f. a & b & c
4. **Please choose the one best answer below. Heated high flow nasal cannula (HFNC) treatment includes the following elements:**
  a. Low flow rate
  b. Precise oxygen concentration delivery
  c. Back-up respiratory rate
  d. Tidal volume delivery
5. **What oxygen flow rate is used for administering <u>low</u> flow oxygen by *<u>nasal cannula</u>*? Select one answer**.
  a. 35 liters per minute
  b. 12 to 15 liters per minute
  c. 6 to 10 liters per minute
  d. 5 liters per minute or lower
6. **Please choose the one best answer below. The normal pH range of arterial blood is:**
  a. 7.15 to 7.25
  b. 7.25 to 7.35
  c. 7.35 to 7.45
  d. 7.45 to 7.55
7. **Please choose the one best answer below. Normal tidal volume value for adults is:**
  a. 10 ml
  b. 100 ml
  c. 500 ml
  d. d. 1,000 ml
8. **Please choose the one best answer below. Lungs that are <u>not</u> compliant are:**
  a. Harder to expand and easier to recoil
  b. Easier to expand and easier to recoil
  c. Harder to expand and harder to recoil
  d. Easier to expand and harder to recoil
9. **Please choose the one best answer below. The Henderson-Hassebach equation for ‘compensation’ is** 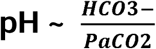. **Another way to think of this equation at the bedside is:**
  a. 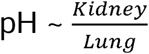
  b. 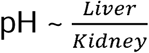
  c. 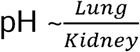
  d. 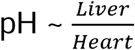
10. **Please choose the one best answer below. The lungs eliminate acid by:**
  a. Reabsorbing HCO_3_-
  b. Exhaling CO_2_
  c. Exhaling H^+^
  d. Excreting H^+^
11. **Please choose the one best answer below. Continuous positive airway pressure (CPAP) treatment includes the following:**
  a. Inspiratory positive airway pressure
  b. Expiratory positive airway pressure
  c. Precise oxygen concentration delivery
  d. Back-up respiratory rate
12. **Please choose the one best answer. The normal pH range of carbon dioxide (CO**_**2**_**) in arterial blood is:**
  a. 35 to 45 mm Hg
  b. 45 to 55 mm Hg
  c. 55 to 65 mm Hg
  d. 65 to 75 mm Hg
13. **Please choose the one best answer below. When pH is low, the lungs respond by:**
  a. Slowing minute ventilation
  b. Decreasing respiratory rate
  c. Decreasing the size of each breath
  d. Increasing tidal volume of each breath
14. **Please choose the one best answer below. The mathematical equation for compliance is:**
  a. *C*/*V*
  b. *V*/*P*
  c. *P*/*V*
  d. *P*/*C*
15. **Please choose the one best answer below. A patient’s arterial blood gas is 7.29 pH / 57 PCO**_**2**_ **/ 29 HCO**_**3**_**-** . **What is the patient’s primary acid base disorder?**
  a. Metabolic acidosis
  b. Respiratory acidosis
  c. Metabolic alkalosis
  d. Respiratory alkalosis
16. **Please choose the one best answer below. A patient’s arterial blood gas is 7.19 pH / 20 PCO**_**2**_ **/ 12 HCO**_**3**_**-**, **and the electrolytes are Na**^**+**^ **139, Cl-117, HCO**_**3**_**-15. What is the patient’s anion gap?**
  a. 5 mEq/L
  b. 10 mEq/L
  c. 7 mEq/L
  d. 12 mEq/L
17. **Please choose the one severity classification of the patient. A 37 year old woman with temperature 40.2 C, cough, respiratory rate of 33 breaths/min, no chest retractions, and oxygen saturation of 91% in room air**.
  a. Mild COVID-19
  b. Moderate COVID-19
  c. Severe COVID-19
  d. Critical COVID-19
18. **Please choose the one best answer below. Intrapleural pressure is:**
  a. Positive pressure between 3 pleural layers
  b. Positive pressure between 2 pleural layers
  c. Negative pressure between 3 pleural layers
  d. Negative pressure between 2 pleural layers
19. **Below are examples of pulse oximeter waveforms. Please select which waveform(s) indicate a quality oxygen saturation measurement. Select one answer**.
  a. 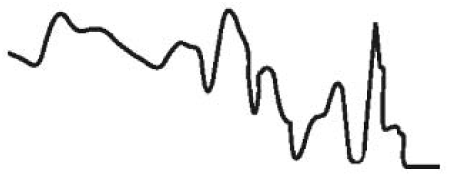
  b. 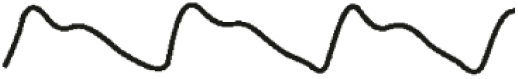
  c. 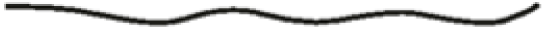
  d. 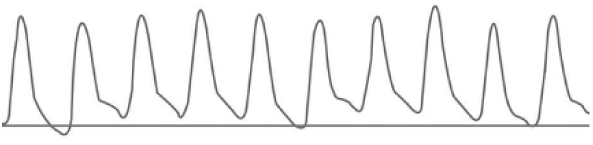
  e. a & d
  f. b & d
  g. a & c
20. **Please choose the one severity classification of the patient. A 65 year old man with temperature 40.2 C, cough, difficulty breathing, respiratory rate of 44 breaths/min, nasal flaring, neck and chest retractions, and oxygen saturation of 96% in room air**.
  a. Mild COVID-19
  b. Moderate COVID-19
  c. Severe COVID-19
  d. Critical COVID-19

## References

1. Zhou P, Yang XL, Wang XG, et al. A pneumonia outbreak associated with a new coronavirus of probable bat origin. Nature 2020;579:270–273.

2. Huang C, Wang Y, Li X, et al. Clinical features of patients infected with 2019 novel coronavirus in Wuhan, China. Lancet 2020;395:497–506.

3. Umakanthan S, Sahu P, Ranade AV, et al. Origin, transmission, diagnosis and management of coronavirus disease 2019 (COVID-19). Postgrad Med J 2020;96:753–758.

4. Johns Hopkins University and Medicine. Coronavirus Resource Center. Available: https://coronavirus.jhu.edu/map.html [Accessed 28 September 2021]

5. United Nations Statistics Division. Lesotho Country Profile. Available: https://data.un.org/CountryProfile.aspx/_Images/CountryProfile.aspx?crName=Lesotho [Accessed 5 September 2021]

6. Avert. Lesotho. Available: https://www.avert.org/professionals/hiv-around-world/sub-saharan-africa/lesotho [Accessed 5 September 2021]

7. endTB. Lesotho. Available: http://www.endtb.org/lesotho [Accessed 5 September 2021]

8. Kruk ME, Gage AD, Arsenault C, et al. High-quality health systems in the Sustainable Development Goals era: time for a revolution. Lancet Glob Health 2018;6:e1196–e1252.

9. Holloway KA, Ivanovska V, Wagner AK, Vialle-Valentin C, Ross-Degnan D. Have we improved use of medicines in developing and transitional countries and do we know how to? Two decades of evidence. Trop Med Int Health 2013;18:656–64.

10. Arsenault C, Jordan K, Lee D, et al. Equity in antenatal care quality: an analysis of 91 national household surveys. Lancet Glob Health 2018;6:e1186–e1195.

11. Jhpiego. Low Dose, High Frequency: A Learning Approach to Improve Health Workforce Competence, Confidence, and Performance. Available: https://hms.jhpiego.org/wp-content/uploads/2016/08/LDHF_briefer.pdf [Accessed 28 September 2021]

12. Evans CL, Johnson P, Bazant E, Bhatnagar N, Zgambo J, Khamis AR. Competency-based training “Helping Mothers Survive: Bleeding after Birth” for providers from central and remote facilities in three countries. Int J Gynaecol Obstet 2014;126:286–90.

13. Enweronu-Laryea C, Engmann C, Osafo A, Bose C. Evaluating the effectiveness of a strategy for teaching neonatal resuscitation in West Africa. Resuscitation 2009;80:1308–11.

14. McCollum ED, Higdon MM, Fancourt NSS, et al. Training physicians in India to interpret pediatric chest radiographs according to World Health Organization research methodology. Pediatr Radiol 2021;51:1322–1331.

15. Horwood C, Butler L, Barker P, et al. A continuous quality improvement intervention to improve the effectiveness of community health workers providing care to mothers and children: a cluster randomised controlled trial in South Africa. Hum Resour Health 2017;15:39.

16. Deorari AK, Paul VK, Singh M, Vidyasagar D. Impact of education and training on neonatal resuscitation practices in 14 teaching hospitals in India. Ann Trop Paediatr 2001;21:29–33.

17. World Health Organization (2005). Handbook: IMCI integrated management of childhood illness. Available: https://apps.who.int/iris/handle/10665/42939 [Accessed 5 September 2021]

18. Word Health Organization (2014). Integrated Management of Childhood Illness: Chart Booklet. Available: https://apps.who.int/iris/bitstream/handle/10665/104772/9789241506823_Chartbook_eng.pdf [Accessed 5 September 2021]

19. Sazawal S, Black RE. Effect of pneumonia case management on mortality in neonates, infants, and preschool children: a meta-analysis of community-based trials. Lancet Infect Dis 2003;3:547–56.

20. Meza PK, Bianco K, Herrarte E, Daniels K. Changing the landscape of obstetric resident education in low-and middle-income countries using simulation-based training. Int J Gynaecol Obstet 2021;154:72–78.

21. Rowe AK, Rowe SY, Peters DH, Holloway KA, Chalker J, Ross-Degnan D. Effectiveness of strategies to improve health-care provider practices in low-income and middle-income countries: a systematic review. Lancet Glob Health 2018;6:e1163–e1175.

